# PTSD symptoms related to COVID-19 as a high risk factor for suicide - Key to prevention

**DOI:** 10.1101/2020.12.15.20246819

**Authors:** Toshinori Chiba, Taiki Oka, Toshitaka Hamamura, Nao Kobayashi, Masaru Honjo, Yuka Miyake, Takatomi Kubo, Hiroyuki Toda, Tetsufumi Kanazawa, Shuken Boku, Akitoyo Hishimoto, Mitsuo Kawato, Aurelio Cortese

## Abstract

**Background:** Rising rates of suicide, the most dreadful consequence of mental health effects elicited by the coronavirus pandemic (COVID-19) are cause for grave concern. However, the exact association between mental health problems and suicide remains largely unknown in relation to COVID-19.

**Methods:** To determine the impact of COVID-19 on suicide trajectory, we used an interrupted time-series design to analyze monthly suicides rates extracted from Japan’s national database. We next used mixed-effects regression models to investigate the relationship between the nationwide suicide increase in August 2020 and psychiatric states of 4,348 individuals from an online survey performed immediately before (December 2019) and during (August 2020) the pandemic. Psychiatric states included depression, anxiety, and COVID-19-related PTSD, a form of severe event-related stress.

**Findings:** In Japan, suicides had gradually decreased before COVID-19 (*β* = −0·7×10^−3^, t_57_ = −14·2, *p* = 8·6×10^−46^), but increased drastically after a state of emergency was declared in April 2020 (β = 0·9×10^−2^, t_57_ = 17·3, *p* = 2·3×10^−67^). We found that PTSD symptoms reliably predict COVID-19’s impact on suicide rates (β = 6·3×10^−4^, t_3936_ = 5·96, *p* = 2·7×10^−9^). In contrast, depression scores are a reliable indicator of stress vulnerability (i.e. future suicide increases, *β* = 0·001, t_3936_ = 6·6, *p* = 4·5×10^−11)^. Simulations revealed that a one-point reduction in PTSD score could decrease suicides by up to 3·1 per ten million people per month in Japan.

**Interpretation:** PTSD symptoms may help to identify high-risk groups so as to increase efficacy of prevention policies.

**Funding:** KDDI collaborative research contract, the Innovative Science and Technology Initiative for Security (JPJ004596), ATLA and AMED (JP20dm0307008).

**Research in context:** *Evidence before this study:* We searched PubMed on December 2, 2020, for “COVID” and “suicid*” in the titles or abstracts of published articles and obtained 269 hits. No language restrictions were applied to the search. Nearly all previous articles on suicide and COVID-19 have reported simulation studies of suicide counts and rates in case studies, editorials, letters, and commentaries. To date, no study has analyzed the association between psychiatric states and suicide increases in the context of the COVID-19 pandemic.

*Added value of this study:* To the best of our knowledge, this is the first study reporting a concrete approach to predict suicide rate increases from psychiatric states during the COVID-19 pandemic. Our findings indicate that PTSD symptoms are a reliable surrogate endpoint of pandemic-related suicide increase.

*Implications of all available evidence:* This work provides a new perspective on preparing guidelines for suicide prevention. Efforts should focus on reducing PTSD severity for single individuals and populations to reduce the overall suicide risk.

## Introduction

The coronavirus (COVID-19) pandemic has affected all aspects of society worldwide, including mental health^1,2^. Many experts have suggested this may lead to an increase in suicides^3–6^. Indeed, on November 30th, CNN reported the shocking news that the suicide rate had increased dramatically in Japan, and that more people died from suicide in October than from COVID in all of 2020^7^. Yet, a recent report in *Lancet Psychiatry* found no suicide increase in Queensland, Australia^8^, where the infection rate is similar to that in Japan (Supplementary Fig 1). Nor has a detectable suicide increase been reported in several countries with high pandemic mortality^3^. But, some have cautioned that suicides may increase later^3,9^. Therefore, suicide prevention in the context of COVID-19 remains a critical public health priority^5,6^. Since country-to-country differences in increased suicides cannot simply be explained by mere infection or mortality rates caused by the pandemic, for effective suicide prevention it is essential to understand the underlying mental/psychiatric mechanisms linking the pandemic to suicides. More importantly, resulting psychiatric states can provide reliable surrogate endpoints for suicide risk. Promising candidates are measures of depression, anxiety, and post-traumatic stress disorder (PTSD)^6,10–12^. However, to date no study has empirically examined associations between these measures and suicide in the context of COVID-19. The objective of this study is to assess the effect of COVID-19 on suicide rates in Japan and to test the reliability of psychiatric states as surrogate endpoints for suicide risk.

**Figure 1.**
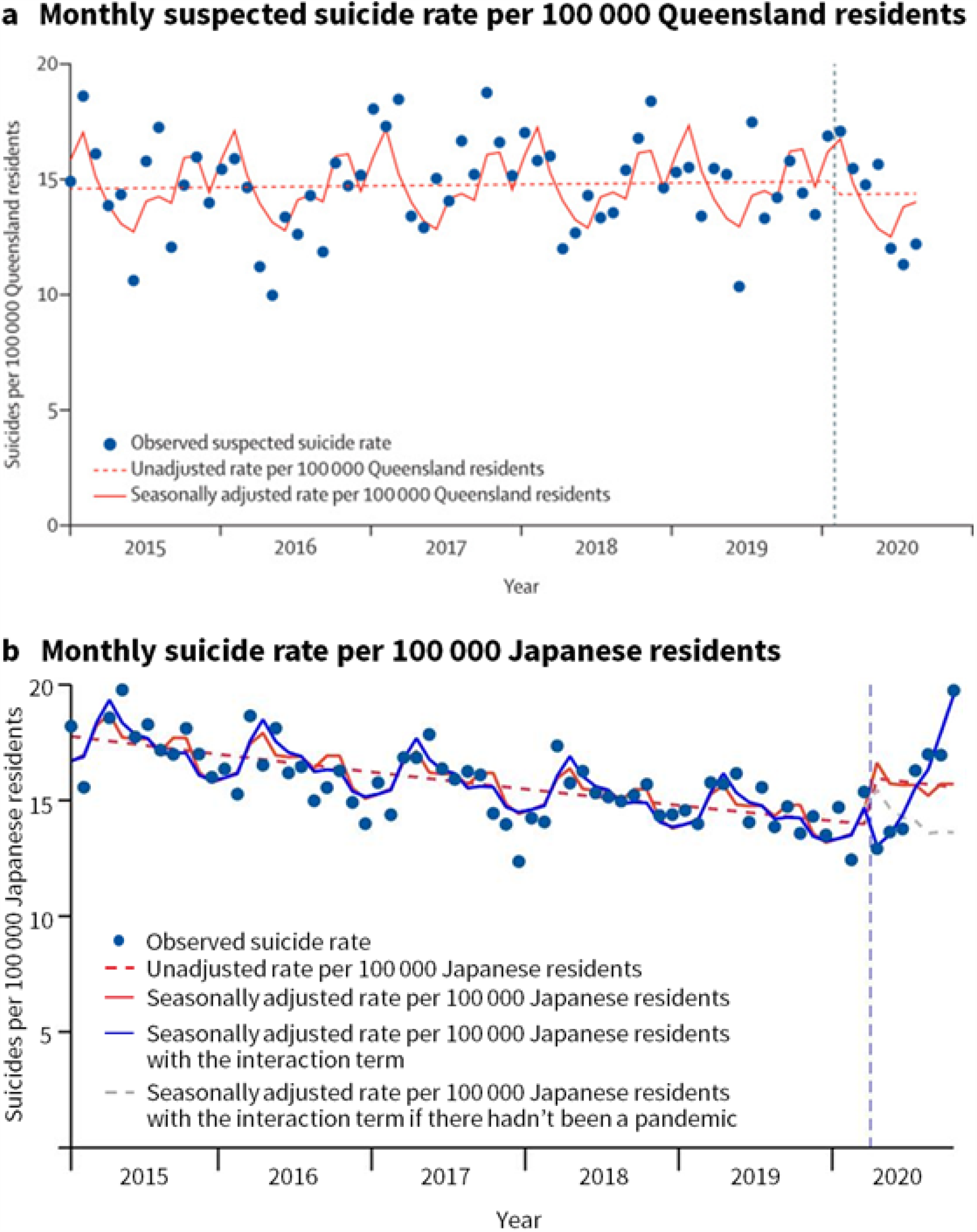
Monthly suicide rates before and during COVID-19. (a) data from Queensland and (b) data from Japan. Because both analyses relied on the latest data, these analyses are based on slightly different time-points (Queensland: April 2015 to August 2020, Japan: April 2015 to October 2020). Nevertheless, the difference between the two trajectories is already apparent in the data until August 2020. Figure 1 (a) is adopted from *Lancet Psychiatry*, November 16, 2020 Online First, Leske et al. “Real-time suicide mortality data from police reports in Queensland, Australia, during the COVID-19 pandemic:an interrupted time-series analysis”, Copyright (2020), with permission from Elsevier.

## METHODS

### Interrupted time-series analyses

Consistent with real-time reports on COVID-19’s impact on suicides in Queensland^8^, we performed interrupted time-series analyses, which have been used to assess the effect of COVID-19 exposure on suicides after accounting for seasonality and pre-exposure non-linear trends. We extracted monthly suicide numbers from January 2015 to October 2020 from the provisional database provided by the Ministry of Health, Labour and Welfare (MHLW) in Japan (https://www.mhlw.go.jp/stf/seisakunitsuite/bunya/0000140901.html, access date: December 2nd). Using these suicide numbers as the outcome, we performed a Poisson regression analysis similar to that performed by Leske *et a*l. on the Queensland data to allow direct comparison^8^. We further examined a model that included the elapsed time since the start of the pandemic to account for cumulative effects (described as “Cumulative” in the model specification). Elapsed time since the onset of the COVID-19 pandemic can be considered an interaction term between the event and time. Under the assumption that pandemic effects do not saturate in the short term, for simplicity we modeled the cumulative effect as linear with respect to time.

Models specification:

mdl1: Unadjusted

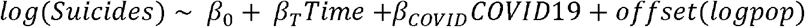

mdl2: Adjusted

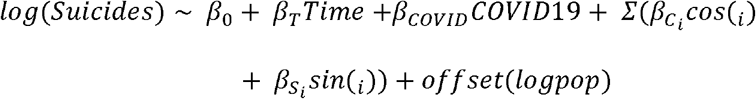

mdl3: Unadjusted with interaction term

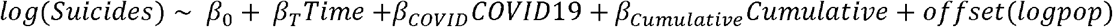

mdl4: Adjusted with interaction term

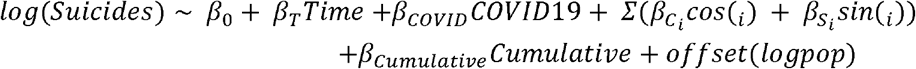

Where *Time* denotes months elapsed since January 2015, *COVID-19* is zero for months before April 2020, and one for months thereafter. Although an absolute definition of the beginning of the pandemic is difficult, we followed Leske *et al*. in defining it as April 2020, when a state of emergency was declared in Japan. *Cos* and *sin* are the Fourier terms consisting of four sine/cosine pairs to adjust for seasonality, with frequency 2pi/12, 2pi/6, 2pi/4, and 2pi/3, respectively. We used the log transformed resident population as an offset variable to transform suicide numbers back to rates.

### Calculation of the increased suicide rate and the proportion attributable to health problems

Using the MHLW database, we calculated the increase in suicide rate from August 2019 to August 2020 for each sex and age group (ten-year bins, e.g., 20-29, etc.). The increase in the suicide rate for each group was defined as follows: the number of suicides in that group in 2019 subtracted from that in 2020, divided by that in 2019.

The MHLW provisional database also provided the general cause of suicides. Up to three choices were allowed for each suicide cause. Categories of causes of suicide were family problems, health (both physical and mental), economy, work, close relationships, school, or others. By dividing the number of suicides due to health problems by the total number of suicides with known causes, we estimated the proportion of the suicide increase explained by health problems.

### Participants and recruitment

This work is part of a larger study on problematic smartphone use, which was approved by the Ethics Committee of the Advanced Telecommunications Research Institute International (Japan). The study was originally planned in 2019, and later expanded to examine the psychiatric impact of COVID-19. Before the COVID-19 pandemic, 99,156 individuals living in the Kansai region of Japan, who registered with an online survey company (Macromill, INC; https://monitor.macromill.com/), were invited via e-mail to participate in screening for the original study, in which they reported their psychiatric status across multiple dimensions. This email contained information about informed consent, and completion of the entire questionnaire was considered to indicate a participant’s consent. Of these, 5,955 individuals were screened and recruited, such that the population evenly included individuals belonging to each quintile in the problematic smartphone use score. In response to the COVID-19 pandemic, volunteers were invited to participate in a follow-up online survey containing additional questions associated with COVID-19. Of these, 4,348 took part in the survey between August 23 and 25, 2020. Furthermore, 198 participants were excluded because there were inconsistencies in their answers. For example, 12 participants answered “yes” to the statement, “I’m not a heavy drinker”, but also “yes” to, “I’m a heavy drinker”. An additional 212 participants were deleted because they responded identically to all items, using only the maximum or minimum value in a questionnaire with reverse scoring. Accordingly, 3,938 responders were included in the current analyses.

### Score measurements

To measure PTSD, depression, and anxiety symptoms, we relied on self-administered questionnaires. The widely used, 22-item Impact of Events Scale-Revised (IES-R)^13^ was employed to assess PTSD symptoms. In our online survey, PTSD symptoms were assessed specifically in regard to COVID-19 (e.g., I thought about COVID-19 when I didn’t mean to). An IES-R score > 32 indicates probable PTSD. To assess depression symptoms, the Center for Epidemiologic Studies Depression Scale (CES-D)^14^ was used. CES-D comprises 20-items, with scores higher than 15 signifying probable depression. The State Trait Anxiety Inventory (STAI-S)^15^ was used to assess state anxiety symptoms. STAI-S comprises 20-items, for which scores higher than 40 or 41 denote probable anxiety disorder for males or females, respectively.

### The association of PTSD severity and increased suicide rate with age or sex

Age and sex effects were removed in the analyses of sex and age, respectively. This was done by mean-subtraction of the effect of sex in the analysis of age-related effects and by regressing the effect of age in the analysis of sex-related effects (taking the residuals of the regression PTSD severity/suicide increase ∼ age). In analyses concerning PTSD severity, all individual data points were considered (N = 3,938), and we used Spearman rank correlation for age and the Mann Whitney U test for sex. In analyses concerning suicide rate increases, data for each age and sex group were used (N = 10), and analyses were weighted by the population size of a given data point (i.e., for each group). We used linear regression for age and logistic regression for sex.

### Simulation of the suicide rate increase using PTSD prevalence of our samples

The suicide rate was simulated based on the prevalence of probable PTSD among our participants, according to the following equation:

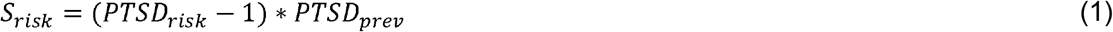

Where *PTSD*_*risk*_ is the risk ratio in suicides (PTSD/Healthy) reported by Fox et al. (2020) (3·96 for males and 6·74 for females), *PTSD*_*prev*_ is the frequency of probable PTSD diagnosis from our samples (i.e., the rate at which the threshold, IES-R > 32, was exceeded).

### Estimation of the effect of a 1-point reduction in IES-R scores on numbers of suicides

We estimated the effect of a 1-point reduction in IES-R scores of all individuals on a simulated suicide rate increase, according to the following equations:

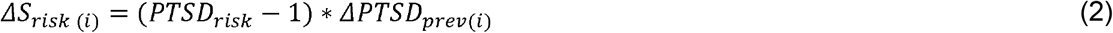

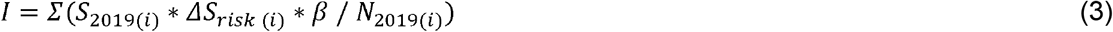

Where *Δ* S_*risk*_ is the difference in the simulated suicide rate increase according to a 1-point reduction in (i) ^th^ sex/age group. *Δ PTSD*_*prev*_ is the difference in the prevalence of probable PTSD diagnosis resulting from a 1-point reduction in the (i) ^th^ group. S_(2019)(*i*)_ is the total number of suicides and N_(2019)(*i*)_) is the population of the (i)^th^ group in August 2019 in Japan. β is the slope of the fitted line between the simulated and the actual suicide increase.

### The possibility of selection bias and the effect of residence

To exclude potential confounding factors due to selection bias, we examined the same linear mixed-effect (LME) model to predict suicide increase from the estimated PTSD severity of each quintile subgroup in the screening population. For this, we adjusted the distribution distortion using a weighted average based on the sample size of each quintile of problematic smartphone use scores. Using PTSD severity estimated from the entire screening population resulted in an outcome (β = 3·8, t_8_ = 4·62, *p* = 0·002) similar to that in the main analysis (β = 6·3×10^−4^, t_3936_ = 5·96, *p* = 2·7×10^−9^).

To test the effect of residence, PTSD severity was compared for each subgroup of participants from Osaka (N = 2,230) with those from locations other than Osaka (N = 1,708). These areas showed a strong correlation in the Pearson correlation analysis (*r* = 0·91, *p* = 2·8×10^−4^). In August 2020, Osaka had the second largest number of people infected with COVID in Japan.

## Results

### Interrupted time-series analyses

All models were superior to the model that assumed an unvarying suicide rate (mdl1: □ ^2^ _66_ = 620, *p* = 2·0×10^−133^, mdl2: □ ^2^ _58_= 810, *p* = 4·4×10^−166^, mdl3: □ ^2^_65_ = 860, *p* = 2·3×10^−185^, mdl4: □ ^2^_66_ =1100, *p* = 3·2×10^−230^). To compare model goodness while controlling for model complexity (i.e., the number of parameters), we used the Bayesian Information Criterion (BIC), with lower values indicating better models. Consistently, mdl4 had the lowest BIC (mdl1: 1430, mdl2: 1280, mdl3: 1190, mdl4: 980). This indicates that the model which accounted for seasonal trends and included the interaction term (i.e., change in the fitted slope before and after the pandemic), best predicted suicide rates. Before COVID-19, suicides had been steadily decreasing (β = −0·7×10^−3^, t_57_ = −14·2, *p* = 8·6×10^−46^), but since the onset of COVID-19 this trend reversed (β = 0·9×10^−2^, t_57_ = 17·3, *p* = 2·3×10^−67^). Additionally, using model mdl4, we estimated the expected suicide rates if there had been no pandemic. We found that actual suicide rates were lower than the estimated rate in the first three months of the pandemic, while they exceeded estimated rates during the following four months (Figure 1).

### Effects of psychiatric states on the increased suicide rate

We utilized psychiatric status data from an online survey (N=3,938 Japanese subjects). Specifically, depression, anxiety, and COVID-19-related PTSD scores were taken during the COVID-19 pandemic. We compared how well these different measures predict the impact of COVID-19 on suicide risk, which was estimated as the increase in suicide frequency from 2019 to 2020 throughout the entire Japanese population (population 125·9 million). Suicide risk was also estimated for different age and sex groups (see Methods).

We used mixed-effects models to test whether psychiatric status (as defined above) could predict the increase in suicide rate. In all models, psychiatric status was considered a fixed effect, while sex was classified as a random effect [model specification: *suicidelncrease ∼* 1 + *psychiatric* + (1 |*sex)*]. We used the BIC to compare model goodness, with smaller values indicating better models. The model based on PTSD scores had the smallest BIC (Figure 2). This indicates that PTSD score is the most effective predictor of increased suicide risk among psychiatric status indicators. To further examine the robustness of PTSD scores as a surrogate endpoint for suicide, we confirmed that adding depression scores and/or anxiety scores to the “main” model (which included only PTSD scores) did not improve the model’s goodness. In these models, PTSD scores were a significant predictor of suicide increase, which indicates that neither the confounder effects of depression nor anxiety can fully explain the association between PTSD and suicide increase. (Figure 2).

**Figure 2.**
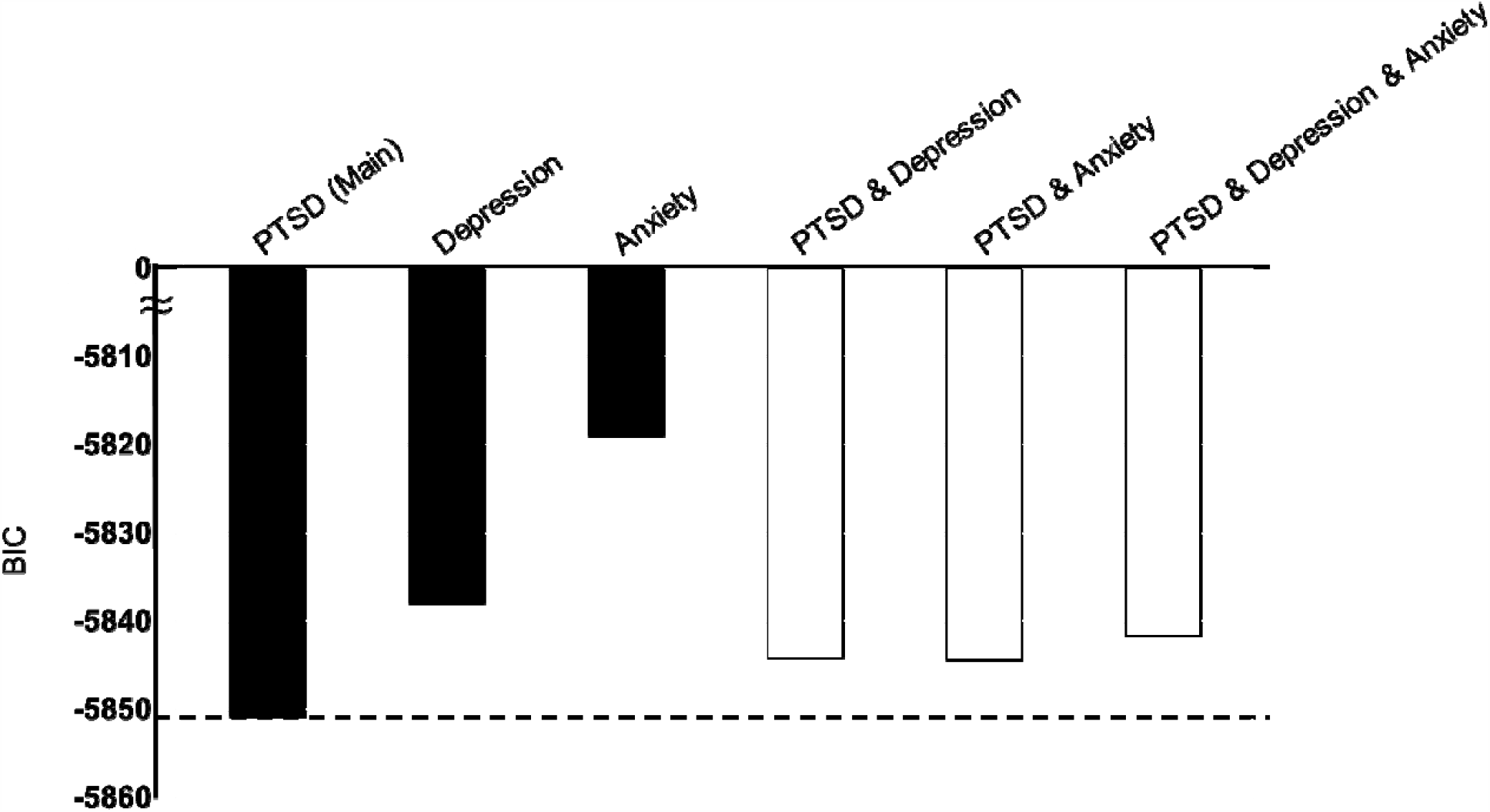
Comparison of models that test associations between psychiatric states and an increased suicide rate. The BIC of each model is presented. Black bars represent models based on PTSD (main model), depression, or anxiety, while white bars represent models based on combinations of PTSD and depression and/or anxiety scores. A model with a bar above the dotted line is worse than the main model.

In the main model, PTSD severity was positively correlated with an increased rate of suicide (*β* = 6·3×10^−4^, t_3936_ = 5·96, *p* = 2·7×10^−9^) (Figure 3a). Next, we examined effects of age and sex on PTSD severity and suicide rate increase, as females and youth were expected to be disproportionately affected by the COVID-19 pandemic^2^. Age was significantly and negatively associated with both increased likelihood of suicide (*β* = −8×10^−3^, t_8_ *= −3*·*05 p* = 0·016) and PTSD severity (N =3938, Spearman’s *rho* = −0·08, *p* = 2·5×10^−7^). Overall, youth appeared more vulnerable to stress, as measured by either PTSD severity or by increased rates of suicide. In contrast, sex was associated only with more frequent suicide (*β* = −1·82, t_8_ *= −2*·*68 p* = 0·028), but not with PTSD severity (N_male_ *=* 2077, N_female_ *=* 1861 *z* = 1·18, *p* = 0·24). Females showed a greater suicide increase than males in all age groups (Figure 3a). To test whether the differential effect of sex on PTSD symptoms and suicide can be explained by previously reported differences in suicide risk ratio associated with PTSD diagnosis, we simulated the suicide increase, based on known risk ratios for each sex^11^ and PTSD prevalence, based on our online survey. A linear regression analysis weighted by the sample size of each data point revealed the strong predictive power of the simulated suicide rate increase (*β* = 0·73, t_8_ = 6·1, *p* = 2·8×10^−4^) (Figure 3b). Thus, the differential suicidal risk-ratio of PTSD thoroughly explained the effect of sex on the suicide increase. Based on this result, we estimated that a one-point reduction of the average PTSD score might lead to a reduction of up to 3·1 suicides per ten million persons per month.

**Figure 3.**
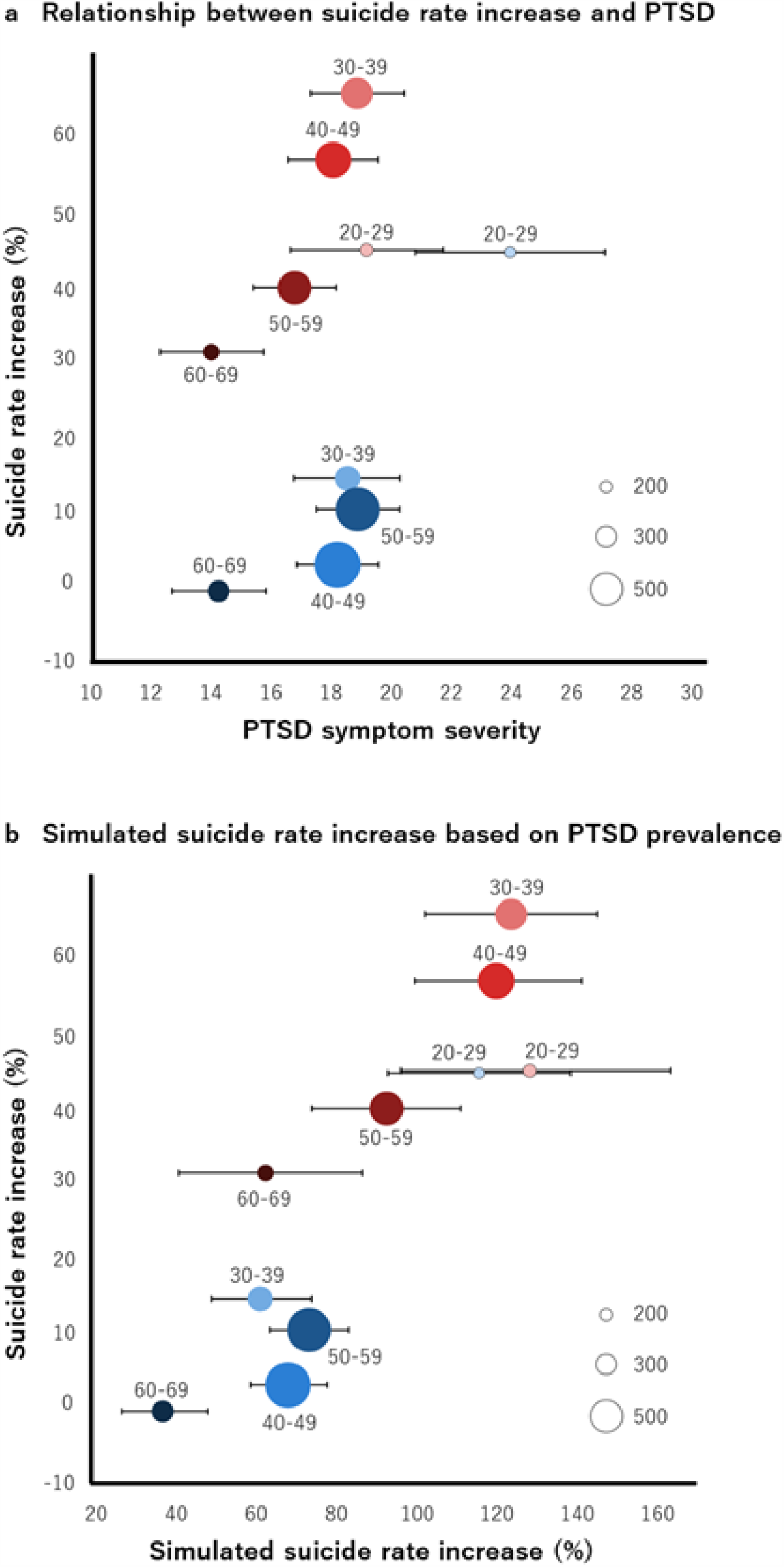
Association between PTSD symptoms related to the COVID-19 pandemic and an increase in the suicide rate. (a) Each circle represents average PTSD severity (X-axis) and the suicide increase (Y-axis) for each age and sex group. (b) Each circle represents the simulated suicide rate increase using PTSD probability (X-axis) and suicide increase (Y-axis) for each age group. Red and blue circles represent female and male data, respectively. The color gradient of each circle represents ages with dark and light colors for older and younger. Corresponding age ranges are described above each circle. The size of each circle represents the number of participants. Circles of various sizes representing 200, 300, and 500 individuals are shown for reference as an inset. Error bars represent bootstrapped 95% confidence intervals.

## Discussion

In contrast to Queensland, Australia, the number of suicides in Japan has been increasing since the declaration of a state of emergency. The increase was particularly noticeable in young and females. The year-on-year suicide rate increase in October reached 2.3 times for females in their twenties. We found that variability in the suicide increase across age and sex groups can be explained by a combination of PTSD symptoms and previously reported sex differences in suicide risk of PTSD. In comparison with other psychiatric symptoms, PTSD symptoms appear to be reliable surrogate endpoints for suicide increase, outperforming depression or anxiety scores in predicting the current suicide increase. We also found that 93·8% (197 out of 210) of additional suicides could be attributed to health (both physical and mental) problems (Supplementary Table1). A substantial proportion of these may be due to PTSD, given its strong association with suicide, as opposed to other mental problems (depression, anxiety). Policies and efforts to reduce PTSD severity within populations, or that of single individuals, are expected to reduce the overall suicide risk. Steps may include promoting an understanding of COVID-19-related PTSD symptoms and of an empirically validated approach for non-experts to mitigate traumatic responses^16^, as well as identifying individuals with high PTSD symptoms, using validated questionnaires in various settings, such as agencies, clinics, and online consultation counters. Individuals identified through these channels should be guided to official clinical settings and given empirically validated treatments focused on PTSD, such as early intervention strategies^17,18^.

To further explore associations between psychiatric states and suicide increase, we also used depression or anxiety scores taken immediately before COVID-19 (see Supplementary). These scores, and particularly depression scores, are strong predictors of both future suicide increases and future PTSD scores. This indicates that depression status is a reliable indicator of stress vulnerability. During the pandemic, special attention should be focused on individuals with a history or current diagnosis of depression. While current depression scores are a reliable indicator of stress vulnerability (future suicide increase), they are not a reliable predictor of current suicide increase in response to COVID-19, for the following reasons. Neither current depression scores nor changes in depression scores from before to during COVID-19 effectively predict the current suicide increase in comparison with PTSD scores (see Supplementary). Therefore, during the pandemic, to prevent suicides, it appears crucial to examine individuals with past or current depression diagnoses from the viewpoint of PTSD scores. Unlike depression or anxiety, PTSD scores may change with increasing suicide. In around April, only 7% of 2697 (medical and non-medical) workers had relatively severe PTSD scores (IES-R > 24) in a Japanese hospital that had already been accepting those infected with COVID-19 at that time (Hishimoto, personal communication). In contrast, in August, we observed that 30.7% of individuals in our online population had relatively severe PTSD scores. This may reflect the increase in year-on-year suicide rates from April (0.88 times) to August (1.3 times). Although further studies of the same population with the same approach are required, increased PTSD scores may serve as a warning sign of potential suicide susceptibility. Furthermore, global surveys with similar approaches, as used here, could help to better explain the reasons behind differences in suicide rate changes across countries, leading to more effective prevention of suicide worldwide. It may be possible to generalize these findings to other large-scale, long-lasting stressful events. The gradual increase in suicides since April, when the state of emergency was declared in Japan, may reflect cumulative stress. In such a scenario, suicides will probably continue to increase until the end of the COVID-19 pandemic. Crucially, this increase may be prevented by PTSD-targeted efforts introduced here.

Since suicide is a rare event, we relied on national data from the Japanese population to reliably estimate the suicide increase, which was compared with our online data. Although the online data may not fully represent the Japanese population, we confirmed that adjusting for selection bias (in our online survey) did not compromise the result. Specifically, by using PTSD severity, estimated from the entire online survey screening population (see Methods), we replicated the original results (*β* = 3·8, t_8_ = 4·62, p = 0·002). Because residence location did not affect PTSD severity within the Kansai region, these results suggest that our online data accurately represent the Japanese population in terms of PTSD severity pattern, according to age and sex. It should be noted that mental illness scores based on online surveys can be higher than those obtained from other types of surveys^19,20^, which may also explain the prevalence of COVID-19-related PTSD diagnosis in our online populations (21·3%), as well as in previous online research (7-35·6%)^21–24^. Consequently, simulated numbers of suicides based on mental illness scores are higher than actual numbers. Although our data suggest the reliability of simulated numbers of suicides as *ratio scales, raw values* are likely to be overestimated. Further, this result, which was based upon Japanese population data, may be of limited application worldwide. Although risk factors for suicide are similar among countries, there are substantial differences in the overall rates of mental disorders among suicides^25^.

In summary, we found that the predictive capacity of PTSD symptoms for the suicide increase during the COVID-19 pandemic exceeded those of depression or anxiety scores. The variability in the suicide increase can be explained by PTSD symptoms across age groups, but not sex. The magnitude of the sex difference in suicide risk was comparable to previously reported sex differences in suicide risk of PTSD. Governments and agencies are encouraged to focus prevention resources on groups at high risk of suicide, identified here: groups showing higher PTSD symptoms.

## Data Availability

The main summary statistics data that support the findings of this study are available within Supplementary Data. Owing to company cohort data sharing restrictions, individual-level data cannot be publicly posted. Data are however available from the authors upon reasonable request and with permission of KDDI Corporation.

## Data sharing

The main summary statistics data that support the findings of this study are available within Supplementary Data. Owing to company cohort data sharing restrictions, individual-level data cannot be publicly posted. However, data are available from the authors upon reasonable request and with permission of KDDI Corporation.

## Corresponding Authors

Toshinori Chiba, MD, Department of Decoded Neurofeedback, Computational Neuroscience Laboratories, Advanced Telecommunications Research Institute International, 2-2-2 Hikaridai, Seika-cho, Soraku-gun, Kyoto 619-0288 JAPAN; Aurelio Cortese, PhD, Department of Decoded Neurofeedback, Computational Neuroscience Laboratories, Advanced Telecommunications Research Institute International.

## Author Contributions

TC and AC made substantial contributions to study conception and design. TC, TO, TH, NK, and MH made substantial contributions to data acquisition. TC, TO, TK and AC conducted statistical analyses. TC, TO, SB, AH, MK, and AC made substantial contributions to interpretation of data. TC, TO and AC drafted the first version of the manuscript. All authors revised and approved the final version of the manuscript. TC, TO and AC take responsibility for the integrity of the work.

## Conflict of Interest Disclosures

This study was funded by KDDI Corporation. There is nothing else to disclose.

## Acknowledgements

We thank Rumi Yorizawa, Misa Murakami, Chika Hosomi, and Miho Nagata for data collection and organization. We also thank Keiko Ide and Takeshi Asami for their helpful discussion.

## Notes

### Author Declarations

We sent participants e-mails contained information about informed consent and completion of the entire questionnaire was considered to indicate a participant's consent. This work is part of a larger study on problematic smartphone use, which was approved by the Ethics Committee of the Advanced Telecommunications Research Institute International (Japan).

## References

1 Holmes EA, O’Connor RC, Perry VH, Tracey I. Multidisciplinary research priorities for the COVID-19 pandemic: a call for action for mental health science. Lancet Psychiatry 2020; 7; 547–60

2 McGinty EE, Presskreischer R, Han H, Barry CL. Psychological distress and loneliness reported by US adults in 2018 and April 2020. JAMA 2020; 324: 93–4.

3 John A, Pirkis J, Gunnell D, Appleby L, Morrissey J. Trends in suicide during the covid-19 pandemic. BMJ 2020; 371: m4352.

4 Wasserman D, Iosue M, Wuestefeld A, Carli V. Adaptation of evidence-based suicide prevention strategies during and after the COVID-19 pandemic. World Psychiatry 2020; 19: 294–306.

5 Gunnell D, Appleby L, Arensman E, et al. Suicide risk and prevention during the COVID-19 pandemic. Lancet Psychiatry 2020; 7: 468–71.

6 McIntyre RS, Lee Y. Preventing suicide in the context of the COVID-19 pandemic. World Psychiatry 2020; 19: 250–1.

7 Wang S, Wright R, Wakatsuki Y. In Japan, more people died from suicide last month than from Covid in all of 2020. And women have been impacted most. CNN 2020; published online Nov 29. https://www.cnn.com/2020/11/28/asia/japan-suicide-women-covid-dst-intl-hnk/index.html (accessed Dec 7, 2020).

8 Leske S, Kõlves K, Crompton D, Arensman E, de Leo D. Real-time suicide mortality data from police reports in Queensland, Australia, during the COVID-19 pandemic: an interrupted time-series analysis. Lancet Psychiatry 2020; published online Nov 16. DOI:10.1016/S2215-0366(20)30435-1.

9 Kõlves K, Kõlves KE, De Leo D. Natural disasters and suicidal behaviours: a systematic literature review. J Affect Disord 2013; 146: 1–14.

10 Blair-West GW, Cantor CH, Mellsop GW, Eyeson-Annan ML. Lifetime suicide risk in major depression: Sex and age determinants. J Affect Disord 1999; 55: 171–8.

11 Fox V, Dalman C, Dal H, Hollander A-C, Kirkbride JB, Pitman A. Suicide risk in people with post-traumatic stress disorder: a cohort study of 3.1 million people in Sweden. J Affect Disord 2020; published online Oct 8. DOI:10.1016/j.jad.2020.10.009.

12 Allgulander C. Suicide and mortality patterns in anxiety neurosis and depressive neurosis. Arch Gen Psychiatry 1994; 51: 708–12.

13 Weiss DS. The Impact of Event Scale: Revised. In: Wilson JP, Tang CS-K, eds. Cross-Cultural Assessment of Psychological Trauma and PTSD. Boston, MA: Springer US, 2007: 219–38.

14 Radloff LS. The CES-D Scale: A Self-Report Depression Scale for Research in the General Population. Appl Psychol Meas 1977; 1: 385–401.

15 Spielberger CD. Manual for the state-trait anxiety inventory (Self-evaluation questionnare). Consulting Psychogyists Press 1970. https://ci.nii.ac.jp/naid/10014833083/ (accessed Nov 16, 2020).

16 Brymer M, Layne C, Jacobs A, Pynoos R, Ruzek J, Steinberg A. Psychological first aid field operations guide (2nd ed.). Los Angeles: National Child Traumatic Stress Network and National Center for PTSD., 2006.

17 Shalev AY, Ankri Y, Israeli-Shalev Y, Peleg T, Adessky R, Freedman S. Prevention of posttraumatic stress disorder by early treatment: results from the Jerusalem Trauma Outreach And Prevention study. Arch Gen Psychiatry 2012; 69: 166–76.

18 Shalev AY, Ankri Y, Gilad M, et al. Long-term outcome of early interventions to prevent posttraumatic stress disorder. J Clin Psychiatry 2016; 77: e580–7.

19 Ballester L, Alayo I, Vilagut G, et al. Accuracy of online survey assessment of mental disorders and suicidal thoughts and behaviors in Spanish university students. Results of the WHO World Mental Health-International College Student initiative. PLoS One 2019; 14: e0221529.

20 Sagar R, Chawla N, Sen MS. Is it correct to estimate mental disorder through online surveys during COVID-19 pandemic? Psychiatry Res 2020; 291: 113251.

21 Forte G, Favieri F, Tambelli R, Casagrande M. COVID-19 Pandemic in the Italian Population: Validation of a Post-Traumatic Stress Disorder Questionnaire and Prevalence of PTSD Symptomatology. Int J Environ Res Public Health 2020; 17. DOI:10.3390/ijerph17114151.

22 Di Crosta A, Palumbo R, Marchetti D, et al. Individual Differences, Economic Stability, and Fear of Contagion as Risk Factors for PTSD Symptoms in the COVID-19 Emergency. Front Psychol 2020; 11: 567367.

23 Karatzias T, Shevlin M, Murphy J, et al. Posttraumatic Stress Symptoms and Associated Comorbidity During the COVID-19 Pandemic in Ireland: A Population-Based Study. J Trauma Stress 2020; published online July 13. DOI:10.1002/jts.22565.

24 Ren Y, Qian W, Li Z, et al. Public mental health under the long-term influence of COVID-19 in China: Geographical and temporal distribution. J Affect Disord 2020; 277: 893–900.

25 Phillips MR, Yang G, Zhang Y, Wang L, Ji H, Zhou M. Risk factors for suicide in China: a national case-control psychological autopsy study. Lancet 2002; 360: 1728–36.

