## Supplementary information for "PTSD symptoms related to COVID-19 as a high risk factor for suicide - Key to prevention"

**Supplementary Table 1.** Causes of suicide in August 2019 and August 2020

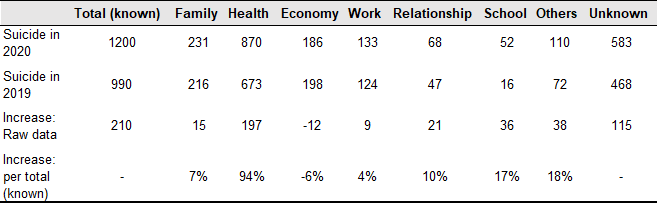

Percentages do not sum to 100 because up to three choices are allowed for each cause.

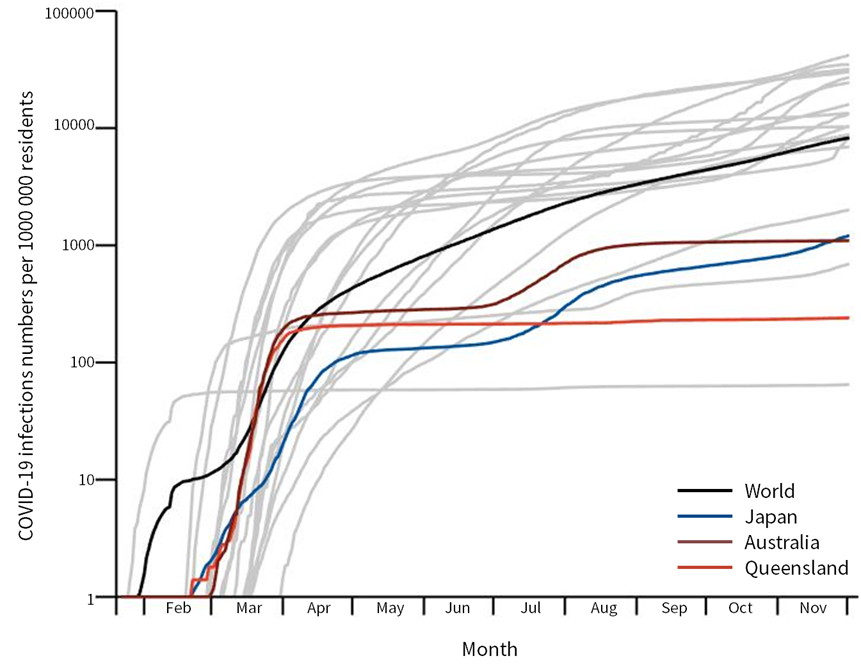

**Supplementary Figure 1.** Cumulative COVID-19 infections per 1 000 000 persons in Japan and Queensland (Australia). Infection rates in G20 countries are shown as gray lines for purposes of comparison.

Data Source:

Queensland government, access date: December 2nd (https://www.qld.gov.au/health/conditions/health-alerts/coronavirus-covid-19/current-status/statistics)

Department of Medical Genome Sciences, Research Institute for Frontier Medicine,

Sapporo Medical University School of Medicine^13^, access date: December 2nd (<https://web.sapmed.ac.jp/canmol/coronavirus/index.html?d=1>)

**Supplementary Texts**

**Effects of pre-COVID-19 psychiatric states on stress response during the pandemic.**

We examined whether depression and anxiety scores reflect stress vulnerability. Specifically, we tested whether “current” psychiatric status can predict “future” PTSD symptoms or a “future” increase in suicide rate using mixed-effect models. Here, psychiatric states include depression and anxiety scores taken before, instead of during, COVID-19. As in the main mixed-effect analyses, psychiatric status was considered a fixed effect, while sex was treated as a random effect. With this model, we first predicted PTSD scores during COVID-19. This revealed that both “current (before COVID-19)” depression and anxiety scores correlated positively with “future (during COVID-19)” PTSD scores (depression: *β* = 0·76, t_3936_ = 33·6, *p* = 1·8 x 10^-217^, anxiety: *β* = 0·60, t_3936_ = 26·3, *p* = 1·4 x10^-140^). Second, we predicted a suicide increase in response to COVID-19 with this model. To allow for a direct comparison with results of main analyses, we compared BICs of these models with that of the “main” model, which predicts a suicide increase based on PTSD scores during COVID-19. As a result, models based on depression, but not on anxiety scores, have smaller BICs in comparison with the main model (Supplementary Figure 2). The model based on depression has a smaller BIC in comparison with that based on anxiety scores. Both “current” depression and anxiety scores were positively correlated with a “future” suicide increase (depression: *β* = 0·001, t_3936_ = 6·6, *p* = 4·5×10^-11^, anxiety: *β* = 0·001, t_3936_ = 6·0, *p* = 2·8 ×10^-9^). These correlations indicate that both depression and anxiety scores are reliable indicators of stress vulnerability, either as measured by future PTSD symptoms or as a suicide increase in response to the COVID-19 pandemic.

**Changes in psychiatric states in response to the pandemic, as predictors of the suicide increase during COVID-19.**

It seemed reasonable that changes in depression or anxiety scores, unlike their current scores, might predict the current suicide increase better than PTSD scores. To examine this possibility, we tested whether changes in depression or anxiety scores from before to during COVID-19 predicted the increase in suicide rate, using mixed-effect models. As in the main mixed-effect analyses, psychiatric status was considered a fixed effect, while sex was treated as a random effect. To allow for direct comparison with results of main analyses, we compared BICs of these models with that of the “main” model, based on PTSD scores during COVID-19. As a result, the main model based on PTSD scores had the smallest BIC (Supplementary Figure 2). This indicates that PTSD score is a more reliable predictor of current suicide risk than changes in depression or anxiety scores.

**Simulation of the increase in suicide rate by depression or anxiety in our study population.**

To further examine the robustness of PTSD scores as a predictor of suicide, we examined whether this linear regression model based on PTSD scores is superior to those based on depression or anxiety scores, as observed in the mixed-effect model. Specifically, we simulated the suicide rate increase based on depression or anxiety scores, instead of PTSD scores, together with their known risk ratios (depression: 12·6 for males and 15·7 for females, anxiety: 6·7 for males and 4·9 for females)^2^^–4^ (Supplementary Figure 3). Then we compared how well these different measures predicted the impact of COVID-19 using BICs. The model based on PTSD scores had the smallest BIC (46·1), compared with those for depression (BIC = 51·8) or anxiety scores (BIC = 53·5). Consistent with the result from the model comparison of main mixed-effects model, this indicates that PTSD score is a better predictor of the increase in suicide risk than depression or anxiety.

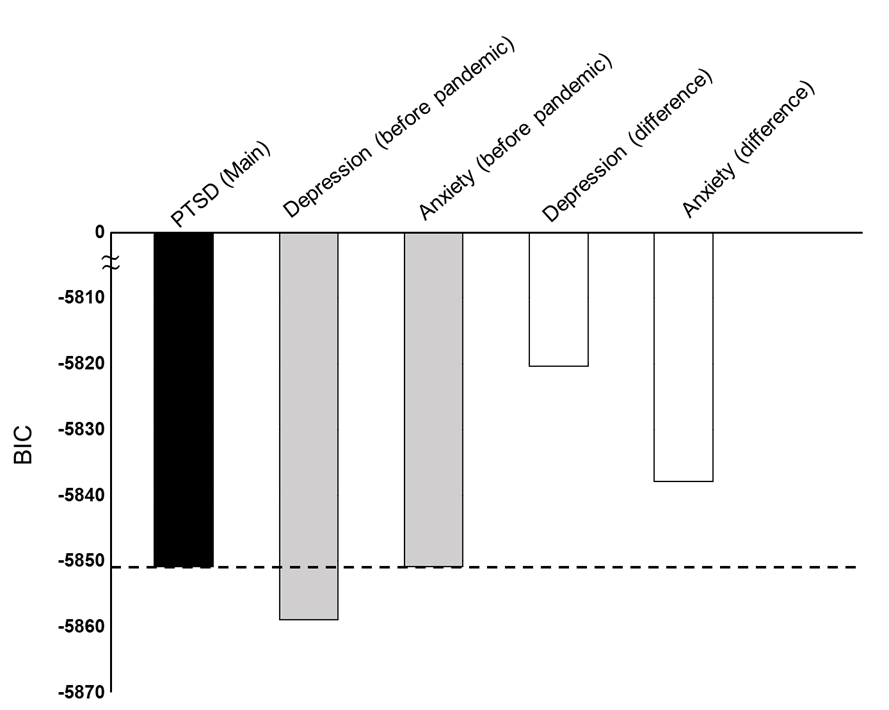

**Supplementary Figure 2.** Comparison of models testing associations between psychiatric states and suicide increases using psychiatric states from before COVID-19. A black bar represents the “main” model based on PTSD symptoms during COVID-19. Gray bars represent models based on depression or anxiety scores before the pandemic. White bars represent models based on changes in depression or anxiety during COVID-19. A model with a bar above the dotted line is a poorer model in comparison with the main model, while a bar below that line is better.

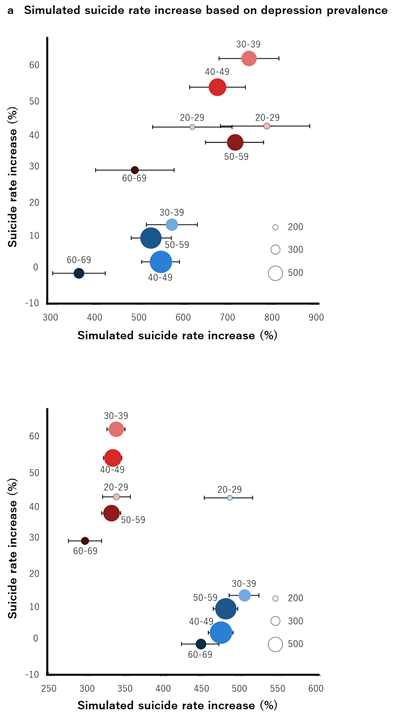

**Supplementary Figure 3.** Association between real suicide rate increase and simulated suicide rate increase by using (a) depression and (b) anxiety probability. Error bars represent bootstrapped 95% confidence intervals.

**Supplementary Data**

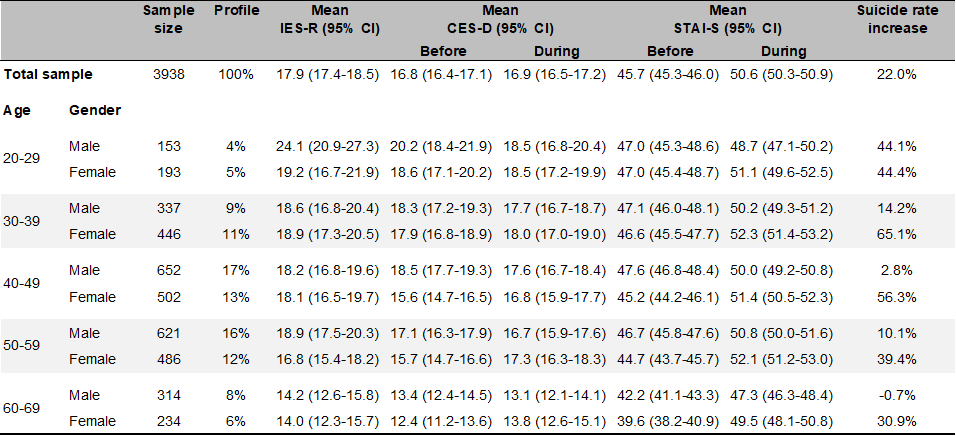

Sample sizes are true (unweighted).

IES-R: Impact of Event Scale-Revised, CES-D: Center for Epidemiologic Studies Depression Scale, STAI-S: State Anxiety Inventory.

Confidence intervals are calculated by using bootstrapped 95%.

**Supplementary Reference**

1	Idogawa M, Tange S, Nakase H, Tokino T. Interactive web-based graphs of novel coronavirus COVID-19 cases and deaths per population by country. *Clin Infect Dis* 2020. <https://academic.oup.com/cid/advance-article-abstract/doi/10.1093/cid/ciaa500/5825754>.

2	Blair-West GW, Cantor CH, Mellsop GW, Eyeson-Annan ML. Lifetime suicide risk in major depression: Sex and age determinants. *J Affect Disord* 1999; **55**: 171–8.

3	Fox V, Dalman C, Dal H, Hollander A-C, Kirkbride JB, Pitman A. Suicide risk in people with post-traumatic stress disorder: a cohort study of 3.1 million people in Sweden. *J Affect Disord* 2020; published online Oct 8. DOI:[10.1016/j.jad.2020.10.009](http://dx.doi.org/10.1016/j.jad.2020.10.009).

4	Allgulander C. Suicide and mortality patterns in anxiety neurosis and depressive neurosis. *Arch Gen Psychiatry* 1994; **51**: 708–12.
